# Clinical application of ISO and CEN/TS standards for liquid biopsies - information everybody wants but nobody wants to pay for

**DOI:** 10.1101/2023.12.04.23299422

**Authors:** Lilli Bonstingl, Christina Skofler, Christine Ulz, Margret Zinnegger, Katja Sallinger, Julia Schönberger, Katharina Schuch, Karin Pankratz, Anatol Borrás-Cherrier, Visnja Somodi, Peter M. Abuja, Lisa Oberauner-Wappis, Thomas Bauernhofer, Thomas Kroneis, Amin El-Heliebi

## Abstract

**Background:** Liquid biopsies are emerging as valuable clinical biomarkers for cancer monitoring. Despite increasing clinical use, standardization remains a challenge. ISO and CEN/TS standardized workflows exist, but their integration into clinical practice is underdeveloped. We aimed to assess the applicability of ISO and CEN/TS liquid biopsy standards in a real-world clinical setting.

**Methods:** We evaluated 659 peripheral blood samples from advanced prostate cancer patients against ISO and CEN/TS standards and tracked all essential criteria. This included assessing tube filing level, complete timing from blood draw until storage, transport conditions, temperature control, hemolysis score and tube draw order and its effects on hemolysis.

**Results:** Among 659 samples, 92.4% (609/659) met the essential criteria for ISO and CEN/TS compliance. In total 83.8% (552/659) of blood collection tubes had high fill levels above 80% of nominal filing level. In our advanced prostate cancer cohort, 12.9% (40/311) of the evaluated plasma samples were hemolytic. Within the draw order of five blood collection tubes, hemolysis did not significantly increase from tube one to five. The complete ccfDNA ISO and CTC CEN/TS workflows were completed within an average of 168 (+/- 71 min) and 248 minutes (+/- 76 min), respectively, from blood draw until storage.

**Conclusions:** Our study demonstrates the feasibility and benefits of adhering to ISO and CEN/T standards in a clinical liquid biopsy study. ISO and CEN/TS standards revealed that hemolysis is a common phenomenon in pre-treated advanced prostate cancer patients, as we eliminated pre-analytical errors as cause.

## Introduction

Liquid biopsies play a crucial role as potentially prognostic and predictive biomarkers and for disease monitoring in cancer (1). While their clinical usage is increasing, standardizations remains a challenging task (2,3). The quality of a blood sample is particularly critical when isolating and analysing low-abundance circulating tumor DNA (ctDNA) fragments or delicate circulating tumor cells (CTCs) (4–6). In routine clinical practice, blood sample collection is prone to various pre-analytical errors, such as inadequate blood collection tubes, underfilling of blood tubes, improper tube inversion, extended transport times, suboptimal temperature conditions during transport and others (7,8). To overcome these challenges, large consortia and societies have developed standards ensuring highest quality of blood samples for subsequent liquid biopsy analysis. These include the European Liquid Biopsy Society (ELBS) (9), CancerID (10), Standardization of generic Pre-analytical procedures for In-vitro DIAgnostics for Personalized Medicine (SPIDIA) (11,12), BLOODPAC (13) and the International Liquid Biopsy Standardization Alliance (ILSA) (14). These efforts have resulted in liquid biopsy pre-analytical standards provided by the International Standards Organisation (ISO) and Technical Specifications from the European Committee for Standardization (CEN/TS). In particular, ISO 20186-3:2019 constitutes a standard for isolation of circulating cell-free DNA (ccfDNA) from plasma, while CEN/TS 17390-3:2020 focuses on specifications for analytical staining of CTCs (15–17). These standards encompass the entire workflow, including documentation, blood sample collection, processing and storage of samples in a controlled environment.

Despite their undeniable importance, widespread implementation of these standards remains limited. The standardisation process requires significant investment in personnel resources for documentation. Only few recent scientific papers dealing with liquid biopsies mention the implementation of these standards (18,19).

In this study, our objective was to evaluate the implementation of ISO and CEN/TS standardized workflows in a real-world clinical liquid biopsy study involving patients with advanced prostate cancer. We aimed to assess the clinical applicability of the standards, to identify relevant pre-analytical parameters and determine common factors leading to non-compliance with ISO or CEN/TS guidelines. The parameters investigated included hemolysis, the filling level and inversion of blood collection tubes, the order of tube draws, transport time and transport conditions.

Here, we present a comprehensive pre-analytical dataset obtained through the systematic application of ISO and CEN/TS standards for ccfDNA and CTCs, which, to our knowledge, has not been reported before. The resulting pre-analytical parameters provide reassurance that samples obtained following these standards meet the quality requirements to justify elaborate and costly liquid biopsy analyses.

## Methods

### Patient cohort

A total of 659 peripheral blood samples were collected from 25 patients of a longitudinal prostate cancer cohort. Patients were diagnosed with castration resistant prostate cancer (CRPC) undergoing change in systemic therapy due to progressive disease. Blood samples for longitudinal liquid biopsy monitoring were collected at the Division of Oncology, Medical University of Graz, Austria, in approximately 12 week intervals with an average number of 5.4 (+/-2.8) visits per patient. At each visit, up to 6 blood tubes were collected.

### Ethic approval

The ethics committee of the Medical University of Graz gave ethical approval for this study protocol and patient information (31-353 ex 18/19) following the declaration of Helsinki and good clinical practice, and written informed consent was obtained from all patients.

### Blood sample collection and processing

Blood samples were collected approximately in 12 week intervals. At each visit, up to 6 blood tubes were collected. The first tube (=tube 0) was not used for ctDNA or CTC detection in this study due to possible contamination with epithelial cells of the skin puncture. The 5 additional blood samples were drawn in the following order: 2 × 10ml PAXgene Blood ccfDNA Tubes (QIAGEN, Hilden, Germany) (each containing 1.5ml cell stabilization additive resulting in a total volume of 11.5ml), followed by 2 × 8.5ml Acid Citrate Dextrose Solution A (ACD-A) tubes (BD, Franklin Lakes, USA) (each containing 1.5ml anti-coagulation and salt solution resulting in a total volume of 10ml) and lastly, an additional 1 × 10ml PAXgene Blood ccfDNA Tube (QIAGEN) (Fig. 1). All samples were collected following the respective ISO and CEN/TS standards. Blood filling levels were assessed for all blood samples.

**Fig. 1:**
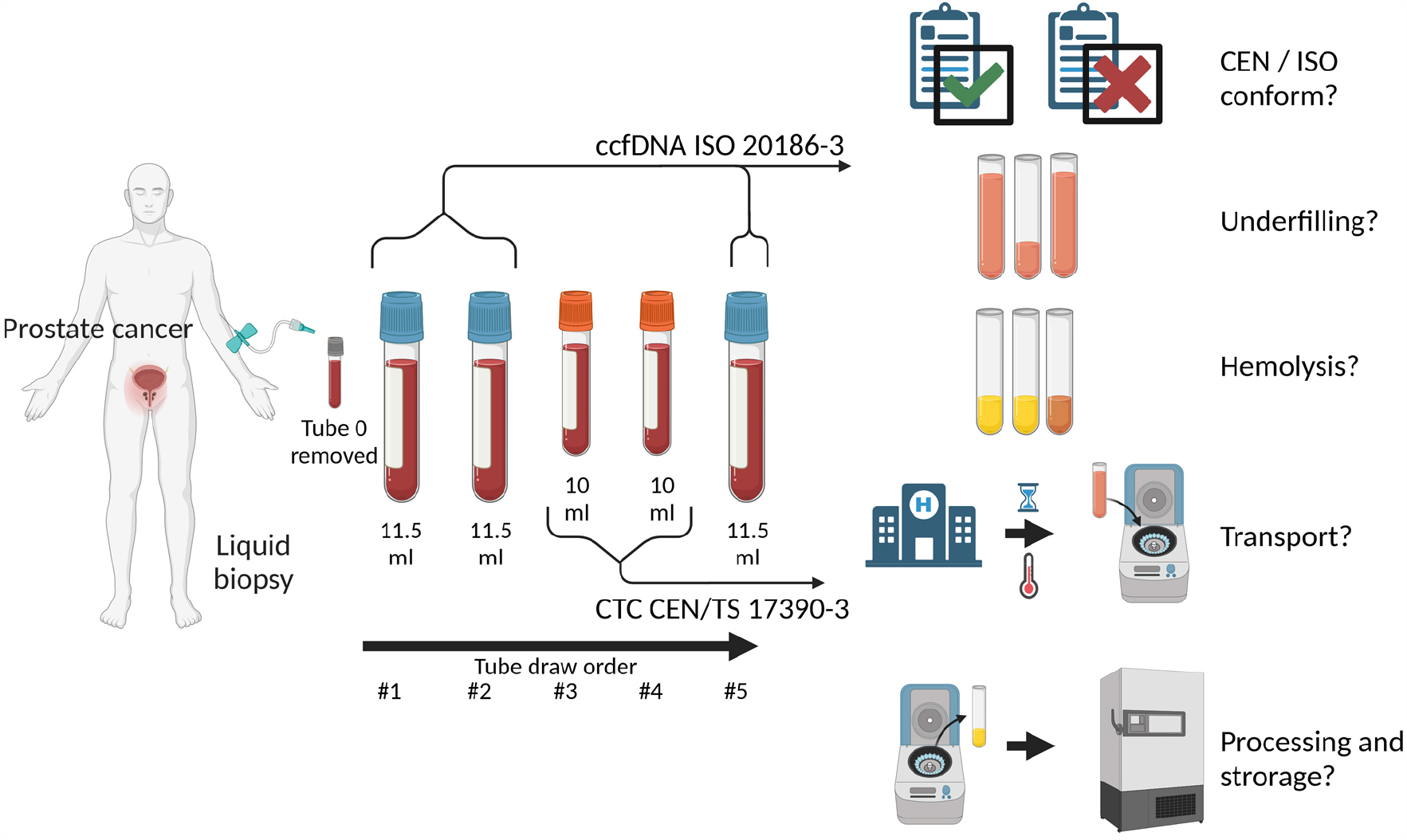
Overview of blood collection tube order, applied ISO CEN/TS standards and parameters to be evaluated.

### ISO and CEN/TS standards

The ISO and CEN/TS standards were followed for processing of PAXgene Blood ccfDNA Tubes (QIAGEN) intended for ctDNA isolation, as outlined in the international standard ISO 20186-3:2019, titled “Molecular in vitro diagnostic examinations – Specifications for pre-examination processes for venous whole blood – Part 3: Isolated circulating cell free DNA from plasma” (16). Similarly, ACD-A tubes intended for CTC isolation and staining were processed according to the Technical Specifications provided by the European Committee for Standardization in CEN/TS 17390-3, titled “Molecular in vitro diagnostic examinations – Specifications for pre-examination processes for circulating tumor cells (CTCs) in venous whole blood – Part 3: Preparations for analytical CTC staining” (17). CTCs were enriched using the Smart Biopsy Cell Isolator (CytoGen, Seoul, South Korea) (20) and the AdnaTest ProstateCancer in accordance with the manufacturer’s instructions (QIAGEN). An overview of the procedures can be found in Fig. 2. The verbal form “shall” in the ISO and CEN/TS standards are “must have” requirements and we assessed the following parameters in detail: patient sample pseudonym ID, date and time of blood drawing, identity of the person who drew blood, identity of the person who processed the blood samples, temperature and storage condition at the blood collection site, verification of tube inversion, temperature conditions during transport, time of call for sample pick-up, time required from blood draw until call, date and time of sample pick-up, date and time of sample arrival at the laboratory, volume of tube fill level, additional notes regarding sample tampering, tube type with catalogue number, lot number, and expiration date, proper labelling of tubes according to standard operating procedures (SOP), centrifugation procedures for tubes, duration from blood draw to processing and storage, timing and storage temperature and location of the sample.

**Fig. 2:**
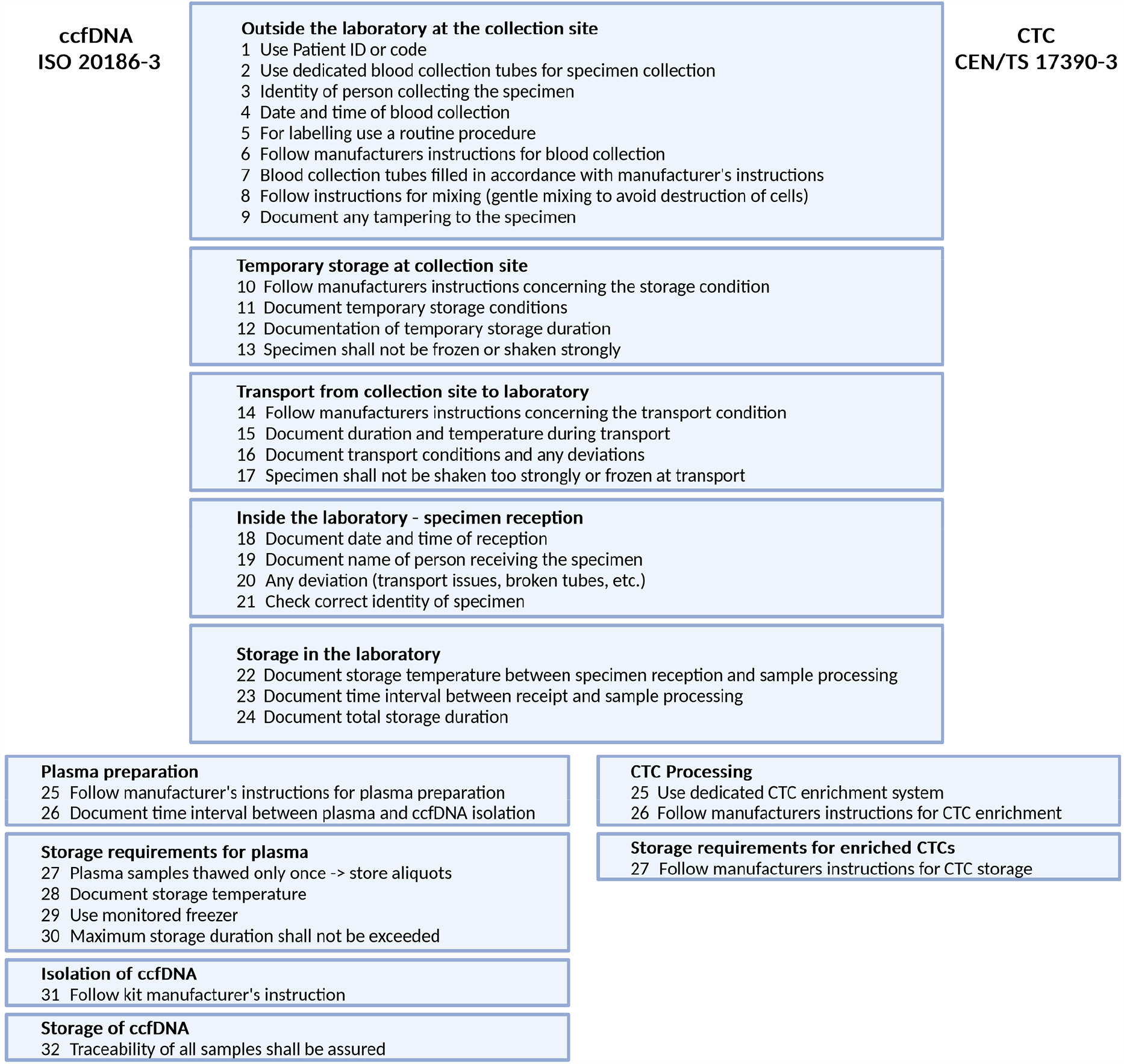
Overview of the criteria, which must be met to assign blood samples as ISO or CEN/TS compliant in our study. Each single step described here is mandatory, omitting or not fulfilling a step will lead to a non-compliant blood sample (16,17).

### Evaluation of hemolysis in plasma samples

Each plasma sample underwent visual and spectrophotometric evaluation to assess red blood cell lysis and lipemia. An aliquot of the plasma samples was analysed using a NanoDrop spectrophotometer (Thermo Fischer Scientific, Waltham, USA). Hemolysis was assessed using a lipemia-independed hemolysis score, as described by Appierto (21). Absorbance was measured at 385nm to identify lipemic samples and 414nm for free haemoglobin. The hemolysis score was calculated using the following formula: Hemolysis score = (Absorbance 414nm-Absorbance 385nm) + 0.1 x Absorbance 385nm (21). Samples with a hemolysis score > 0.25 were classified as hemolytic as described previously (22).

## Results

### Compliance with ISO and CEN/TS standards

Of the total 659 blood samples in our clinical cohort, 92.4% (609/659) met the essential criteria to be considered ISO and CEN/TS compliant. The remaining 7.6% (50/659) of samples did not meet ISO and CEN/TS compliance due to missing information regarding the identity of the person who collected the blood specimen (6.8%, 45/659) and lack of tube inversion after blood collection (0.8%, 5/659).

### Tube fill level assessment

We assessed the fill level of both PAXgene ccfDNA tubes (11.5ml total volume, including tube supplements) and ACD-A tubes (10ml total volume, including tube supplements). The mean fill level of all 659 blood tubes was 92.4% +/- 15.8 (Fig. 3A). The mean fill level for tube 1 was 88.9% +/- 16.7 (N=133), tube 2: 87.8% +/- 18.2 (N=133), tube 3: 97.6% +/- 12.8 (N=132), tube 4: 98.4% +/- 10.0 (N=130) and tube 5: 89.6% +/- 16.5 (N=131).

**Fig. 3:**
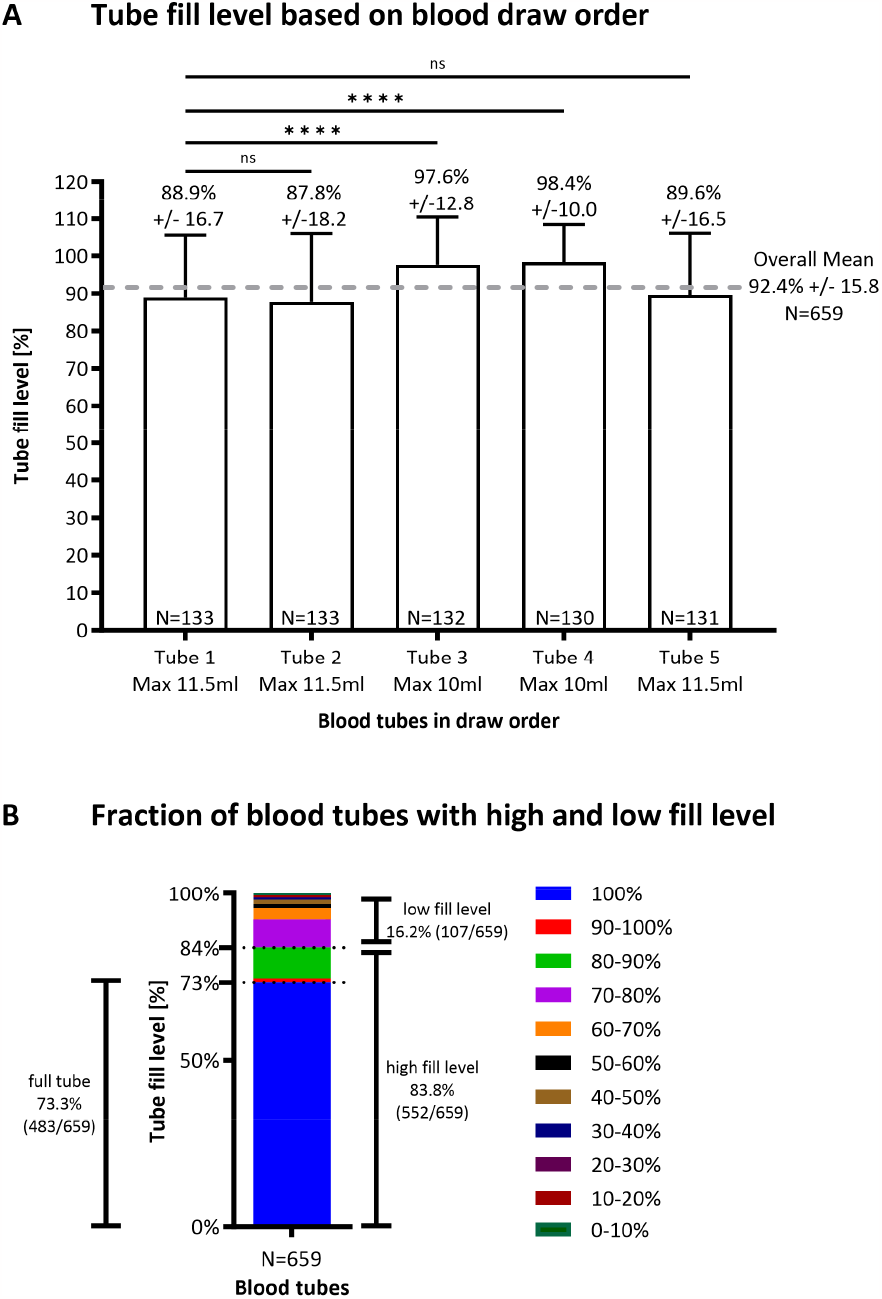
Tube fill level assessment. A) Evaluation of the level of tube fullness based on draw order. B) Fraction of blood collection tubes with high and low fill level. **** p< 0.0001, one-way ANOVA, ns = not significant.

Notably, we observed differences between the two tube types, with the 11.5ml PAXgene ccfDNA tubes exhibiting a lower mean fullness level compared to the 10ml ACD-A tubes, measuring 88.8% +/- 17.1 (N=397) versus 98.0% +/-11.5 (N=262), respectively (p< 0.0001, one-way ANOVA). Interestingly, there was no statistically significant difference between the first and the last tube drawn from each patient (p= 0.6917, one-way ANOVA). To further categorize the filling level performance, we divided it into 10% steps. We defined fill levels >80% as high and those ranging from 1% to 79% as low. Of the total 659 blood collection tubes evaluated, 83.8% (552/659) had a high fill level, while the remaining 16.2% (107/659) had a low fill level (Fig. 3B). Of all tubes, 73.3% (483/659) were completely filled.

### Hemolysis score

Of the 659 blood samples, 311 samples were forwarded to plasma isolation including assessment of the hemolysis score for this study. Among the 311 plasma samples, the majority, 87.1% (271/311), were classified as non-hemolytic, with a hemolysis score of lower than 0.25. In contrast, the remaining 12.9% (40/311) exhibited hemolysis, with a hemolysis score above 0.25 (Fig. 4A). When examining if there are statistically more or less hemolytic samples in the first, second or last blood draw, we did not observe a significant difference related to the blood draw order (p= 0.4626 one-way ANOVA). We discovered that 17.3% (22/127) of samples from tube 1, 7.4% (7/94) of samples from tube 2 and 12.2% (11/90) of samples from tube 5 displayed hemolysis (hemolysis score >0.25).

**Fig. 4:**
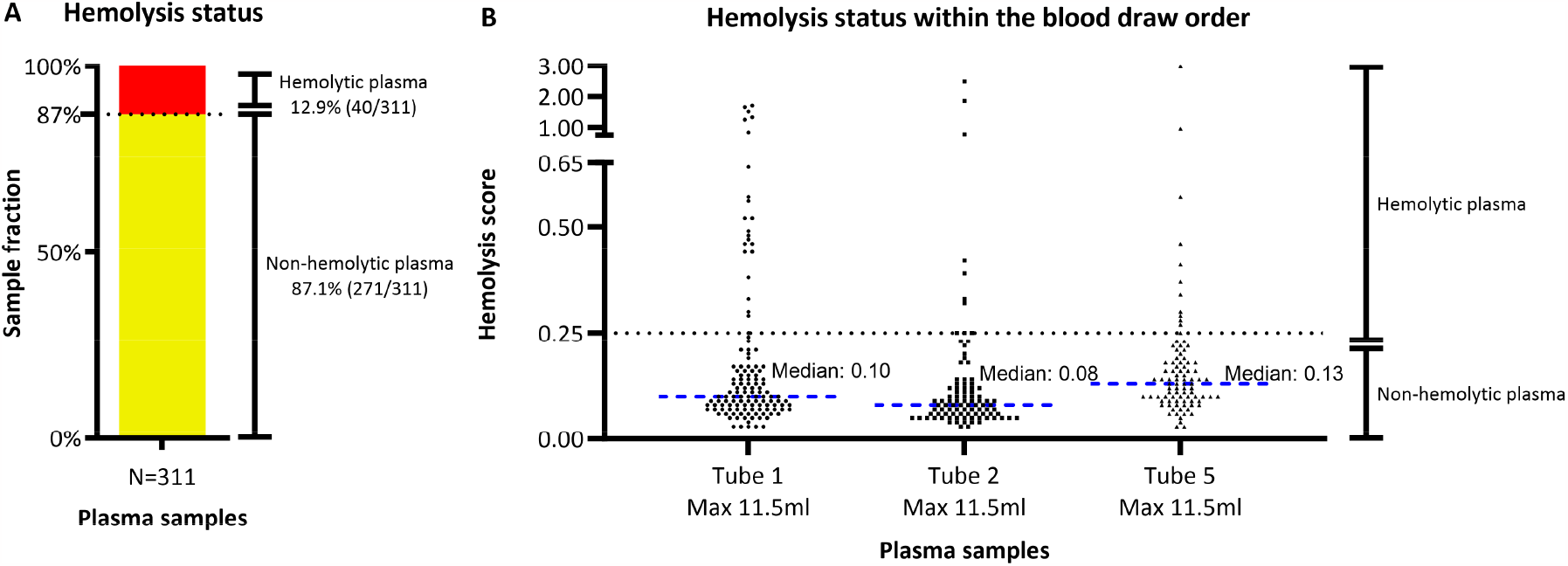
Hemolysis status of plasma samples based on the spectroscopically measured hemolysis score. A) Fraction of hemolytic plasma samples. B) Hemolysis assessment within the tube order, with no statistically significant difference (p= 0.4626 one-way ANOVA). Dashed line indicates hemolysis score threshold of 0.25.

### Blood tube order and hemolysis

We aimed to investigate whether hemolysis levels differed among multiple blood tubes collected at the same sampling time point. For instance, we sought to determine if the presence of hemolysis in the first blood collection tube would indicate consistent hemolysis in subsequent tubes. Therefore, we assessed 121 blood draw time points, where a minimum of two and a maximum of six blood collection tubes were obtained at each time point. We assessed the hemolysis score only for tubes where plasma was isolated (tubes 1, 2 and 5). The remaining tubes were directly forwarded to CTC isolation (tube 3 and 4) without plasma isolation, or were removed due to possible skin cell contamination (tube 0). Our findings revealed that in 77.7% (94/121) of a blood draw series, the plasma remained non-hemolytic for all tubes (Fig. 5). In 5.8% (7/121) of the blood draw series, all blood tubes showed hemolysis. Interestingly, in 11.6% (14/121) of the blood draw series, only the first tube showed hemolysis, but subsequent tubes were not hemolytic. In contrast, 5.0% (6/121) of the blood draw series exhibited initial tubes without hemolysis, but subsequent tubes showed detectable hemolysis (Fig. 5). Notably, we did not observe any instances of a transition from hemolytic to non-hemolytic and then back to hemolytic, nor vice versa, within any of the blood draw series.

**Fig. 5:**
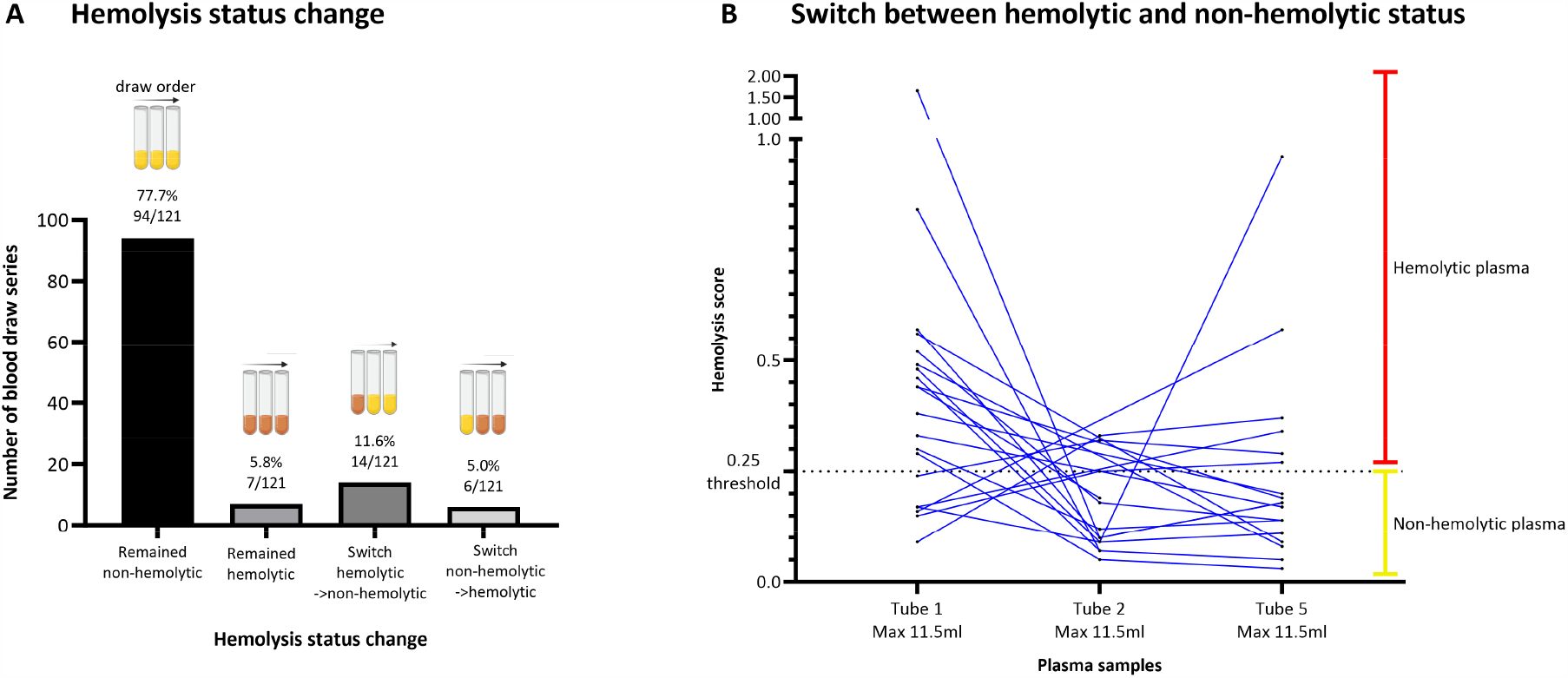
Hemolysis status on sequentially drawn blood samples. A) Hemolysis changes during the draw order. B) Detailed overview of the hemolysis score of plasma samples, which switched from hemolytic to non-hemolytic status and vice versa. Samples, which did not switch their hemolysis status, are not shown for better clarity.

### Timing of samples

Thorough documentation enabled establishing a time line for each blood sample and keeping track of its condition. In our clinical setting, the mean time from blood sampling to pick-up for transport was 32 +/- 27 minutes (Fig. 6). The average duration from blood draw to arrival at the laboratory was 45 +/- 29 minutes, after which the samples were forwarded to CTC isolation or plasma isolation. The mean time from blood draw to start of CTC isolation was 62 +/- 37 minutes. For plasma isolation and storage, adhering to the ISO 20186-3:2019 standards, the full process was completed within a mean time of 168 +/- 71 minutes. Regarding CEN/TS 17390-3:2020 compliant CTC isolation and storage for subsequent staining procedures, the process was completed within a mean of 248 +/- 60 minutes.

**Fig. 6:**
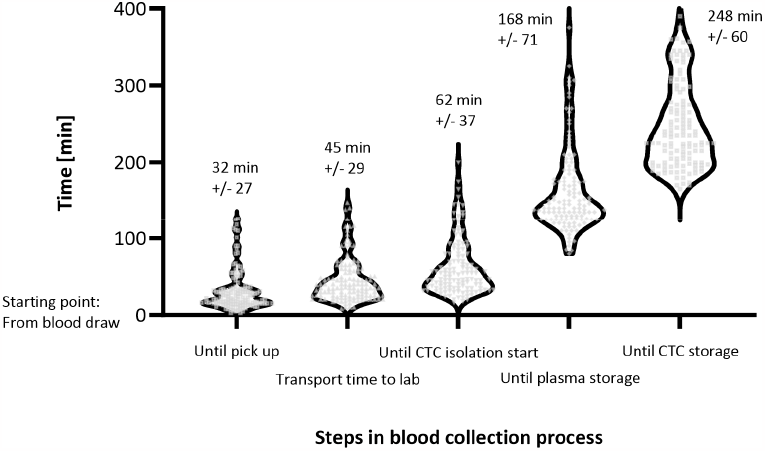
Documented timeline for each sample following the whole ISO and CEN/TS workflows. Timeline represented in minutes, values above the violin plot are mean values with standard deviation.

## Discussion

Our study demonstrates the feasibility and benefits of adhering to ISO and CEN/TS standards in a clinical liquid biopsy study. We found that over 92% of the blood samples in our study met the criteria for ISO and CEN/TS compliance. The most frequently missed criterion was the documentation of the identity of the person who collected the blood specimen. Only a small fraction of samples failed to comply with the standards due to missing tube inversion. Notably, our results indicate that hemolysis is a common phenomenon in advanced prostate cancer patients, which is likely associated with patient-specific factors such as therapy or disease progression, as we can rule out pre-analytical errors as causative factor.

By following the ISO and CEN/TS standards and ensuring thorough documentation, we effectively eliminated pre-analytical errors, such as missing tube inversion, prolonged times of sample processing, or unknown exposure to temperature variations (23). This enabled us to investigate hemolysis in advanced prostate cancer samples and identified increased levels in approximately 13% of all samples. Indeed, hemolysis analysis can only be properly addressed if the pre-analytical parameters are strictly defined and followed. Otherwise, hemolysis could be caused by multiple other pre-analytical factors than the patient-specific effects. Surprisingly, we observed only a minimal increase in hemolysis whenever multiple tubes were collected in one session, despite the expectation that a higher number of blood collection tubes would increase the probability of hemolysis due to prolonged tourniquet application (24).

In our study, with six blood tubes collected sequentially, and the first tube removed due to possible contamination with skin cells, we found that blood tubes drawn later in the collection process had no statistically difference in the hemolysis score (Fig. 4). However, we observed that in 11.6% of our blood draw series, the first tube was hemolytic and the following were non-hemolytic (Fig. 5A). These findings suggest that later tubes may be preferable for subsequent liquid biopsy analysis. Although we did not specifically investigate the impact of hemolysis on follow-up ctDNA or CTC analysis, previous studies have shown that hemolysis may negatively affect liquid biopsy analysis, leading to lower ctDNA yield or issues with blood coagulation during CTC enrichment (25–28). Another interesting finding from our hemolysis analysis was that a switch from non-hemolytic to hemolytic status of the sample (11.6%), and vice versa (5.0%) is a quite frequent phenomenon. If it happened, it only occurred once in our series of six blood samples. For example, if the first tube was non-hemolytic, the subsequent second tubes could become hemolytic (= one switch), with all remaining tubes also displaying hemolytic status. In our study, we did not observe a dual switch, for instance, from non-hemolytic to hemolytic and then back to non-hemolytic status.

We also investigated the tube fill levels in the consecutively drawn tubes. We did not observe a statistically significant difference between tubes 1 and 5, despite expecting a decrease in fill level with each subsequent tube due to possible prolonged venous stasis by long tourniquet placement (29). Additionally, more than 83% of blood tubes displayed high fill levels. An important conclusion is that special attention is needed when filling high volume tubes, such as PAXgene ccfDNA tubes. While their caps are equipped with spill-over protection to ensure secure opening of the blood tubes, this safety feature poses a challenge for visually assessing the fill level during blood collection. This becomes particularly relevant for high volume tubes, especially when drawing the last millilitre of blood into an almost full tube. As a result, the mean fill level of these larger tubes was lower (approximately 89%) compared to smaller tubes like ACD-A (approximately 98%). Despite the lower fill level, both tube types provided sufficient material for follow-up analyses, with the larger tubes containing stabilizing agents to ensure high-quality ccfDNA for subsequent analysis.

In our study, we meticulously tracked the timing of every step involved in blood collection, transport, and processing, in accordance with the demands of the ISO and CEN/TS standards. Our laboratory implemented a fast turnaround time of approximately 2.5 hours for ISO-compliant plasma storage for ccfDNA extraction and roughly 4 hours for CEN/TS-compliant CTC enrichment and storage. These processing times comprise the entire workflow, from the moment of blood draw to final storage. The personnel resources required for managing and processing involved two full-time equivalents of biomedical scientists serving as liquid biopsy managers. Notably, we successfully addressed a pivotal challenge – the elapsed time between blood draw and sample pick-up. This interval is often an unknown variable and difficult to control in many liquid biopsy studies where blood samples are collected during clinical routine and forwarded for research purposes. Factors like temperature variations, tube inversion and the need for pseudonymization further complicates these challenges. Our observations revealed that the presence of dedicated liquid biopsy managers greatly enhanced our ability to adhere to ISO and CEN/TS standards in a clinical setting.

The application of ISO and CEN/TS standards in liquid biopsy samples offers a significant advantage to biobanks, leveraging their liquid specimens for future projects. This has also been identified as an important factor by European research infrastructure for biobanking (https://www.bbmri-eric.eu) (30). The pre-analytical data collected becomes pivotal for development of novel liquid biopsy assays, seeking to access previously collected liquid biopsy cohorts within biobanks. ISO and CEN/TS conform biobanked liquid biopsy samples may reduce the necessity for new prospective clinical studies, allowing a more time-efficient and cost-effective development of liquid biopsy assays. Similarly, within the in vitro diagnostic regulation (IVDR) in the European Union and the US Food and Drug Administration (FDA), pre-analytical parameters are becoming crucial for liquid biopsy workflows (31).

In conclusion, our study underscores the invaluable insights that are obtained by adhering to the available ISO and CEN standards for liquid biopsies. Indeed, this is “information everybody wants but nobody wants to pay for” to quote Dr. Howard I Scher from the Memorial Sloan Kettering Cancer Center, New York, USA. Although implementation of ISO and CEN/TS standards may require substantial investment in terms of personnel and resources, it provides information on critical pre-analytical parameters and ensures that costly and elaborate liquid biopsy analyses are built on a solid foundation.

## Data Availability

All data produced in the present work are contained in the manuscript

## Data availability

All data produced in the present study are contained in the manuscript.

## Acknowledgements

The authors thank Daniel Kummer for technical support and Gerlinde Gornicec, Carina Kreuter, Sylvia Tripolt, Karin Groller and Lisa Jaritz from the study coordination team of the Division of Oncology, Medical University of Graz for their excellent support in patient enrolment. Figures were created with BioRender.com and Prism GraphPad.

## References

1. Alix-Panabières C, Pantel K. Liquid Biopsy: From Discovery to Clinical Application. Cancer Discov. 2021 Apr;11(4):858–73.

2. Heitzer E, van den Broek D, Denis MG, Hofman P, Hubank M, Mouliere F, et al. Recommendations for a practical implementation of circulating tumor DNA mutation testing in metastatic non-small-cell lung cancer. ESMO Open. 2022 Apr;7(2):100399.

3. Pascual J, Attard G, Bidard FC, Curigliano G, De Mattos-Arruda L, Diehn M, et al. ESMO recommendations on the use of circulating tumour DNA assays for patients with cancer: a report from the ESMO Precision Medicine Working Group. Ann Oncol. 2022 Aug;33(8):750–68.

4. El-Heliebi A, Hille C, Laxman N, Svedlund J, Haudum C, Ercan E, et al. In Situ Detection and Quantification of AR-V7, AR-FL, PSA, and KRAS Point Mutations in Circulating Tumor Cells. Clinical Chemistry. 2018 Mar;64(3):536–46.

5. El-Heliebi A, Heitzer E. State of the Art and Future Direction for the Analysis of Cell-Free Circulating DNA. In: Nucleic Acid Nanotheranostics [Internet]. Elsevier; 2019 [cited 2021 Mar 7]. p. 133–88. Available from: https://linkinghub.elsevier.com/retrieve/pii/B9780128144701000058

6. Hofmann L, Sallinger K, Haudum C, Smolle M, Heitzer E, Moser T, et al. A Multi-Analyte Approach for Improved Sensitivity of Liquid Biopsies in Prostate Cancer. Cancers (Basel). 2020 Aug 11;12(8).

7. Bonini P, Plebani M, Ceriotti F, Rubboli F. Errors in laboratory medicine. Clin Chem. 2002 May;48(5):691–8.

8. Lippi G, von Meyer A, Cadamuro J, Simundic AM. Blood sample quality. Diagnosis (Berl). 2019 Mar 26;6(1):25–31.

9. UKE - ELBS – European Liquid Biopsy Society [Internet]. [cited 2023 Oct 2]. Available from: https://www.elbs.eu

10. IMI Innovative Medicines Initiative [Internet]. 2015 [cited 2023 Oct 2].IMI Innovative Medicines Initiative | CANCER-ID | Cancer treatment and monitoring through identification of circulating tumour cells and tumour related nucleic acids in blood. Available from: http://www.imi.europa.eu/projects-results/project-factsheets/cancer-id

11. Oelmueller U. Standardization of generic pre-analytical procedures for in vitro diagnostics for personalized medicine. N Biotechnol. 2021 Jan 25;60:1.

12. Spidia [Internet]. [cited 2023 Oct 2]. Available from: https://www.spidia.eu/

13. BLOODPAC [Internet]. 2023 [cited 2023 Oct 2]. BLOODPAC. Available from: https://www.bloodpac.org

14. Connors D, Allen J, Alvarez JD, Boyle J, Cristofanilli M, Hiller C, et al. International liquid biopsy standardization alliance white paper. Crit Rev Oncol Hematol. 2020 Dec;156:103112.

15. Dagher G, Becker KF, Bonin S, Foy C, Gelmini S, Kubista M, et al. Pre-analytical processes in medical diagnostics: New regulatory requirements and standards. N Biotechnol. 2019 Sep 25;52:121–5.

16. Standards I. ISO. [cited 2023 Oct 2]. ISO 20186-3:2019. Available from: https://www.iso.org/standard/69800.html

17. Standards C. ONR CEN/TS 17390-3: 2020 05 01 - Molecular in vitro diagnostic examinations - Specifications for pre-examination processes for circulating tumor cells (CTCs) in venous whole blood - Part 3: Preparations for analytical CTC staining (CEN/TS 17390-3:2020) [Internet]. 2020 [cited 2023 Oct 2]. Available from: https://shop.austrian-standards.at/action/en/public/details/675558/ONR_CEN_TS_17390-3_2020_05_01

18. Mordenti M, Capicchioni V, Corsini S, Locatelli M, Abelli E, Banchelli F, et al. Preanalytical DNA assessment for downstream applications: How to optimize the management of human biospecimens to support molecular diagnosis-An experimental study. J Clin Lab Anal. 2022 Jul;36(7):e24531.

19. Salvianti F, Gelmini S, Mancini I, Pazzagli M, Pillozzi S, Giommoni E, et al. Circulating tumour cells and cell-free DNA as a prognostic factor in metastatic colorectal cancer: the OMITERC prospective study. Br J Cancer. 2021 Jul;125(1):94–100.

20. Choi SY, Lim B, Kyung YS, Kim Y, Kim BM, Jeon BH, et al. Circulating Tumor Cell Counts in Patients With Localized Prostate Cancer Including Those Under Active Surveillance. In Vivo. 2019;33(5):1615–20.

21. Appierto V, Callari M, Cavadini E, Morelli D, Daidone MG, Tiberio P. A lipemia-independent NanoDrop(®)-based score to identify hemolysis in plasma and serum samples. Bioanalysis. 2014 May;6(9):1215–26.

22. Zhelankin AV, Stonogina DA, Vasiliev SV, Babalyan KA, Sharova EI, Doludin YV, et al. Circulating Extracellular miRNA Analysis in Patients with Stable CAD and Acute Coronary Syndromes. Biomolecules. 2021 Jun 29;11(7):962.

23. Magnette A, Chatelain M, Chatelain B, Ten Cate H, Mullier F. Pre-analytical issues in the haemostasis laboratory: guidance for the clinical laboratories. Thromb J. 2016;14:49.

24. Saleem S, Mani V, Chadwick MA, Creanor S, Ayling RM. A prospective study of causes of haemolysis during venepuncture: tourniquet time should be kept to a minimum. Ann Clin Biochem. 2009 May;46(Pt 3):244–6.

25. Nishimura F, Uno N, Chiang PC, Kaku N, Morinaga Y, Hasegawa H, et al. The Effect of In Vitro Hemolysis on Measurement of Cell-Free DNA. J Appl Lab Med. 2019 Sep;4(2):235–40.

26. Pizzamiglio S, Zanutto S, Ciniselli CM, Belfiore A, Bottelli S, Gariboldi M, et al. A methodological procedure for evaluating the impact of hemolysis on circulating microRNAs. Oncol Lett. 2017 Jan;13(1):315–20.

27. Shin S, Woo HI, Kim JW M D YK, Lee KA. Clinical Practice Guidelines for Pre-Analytical Procedures of Plasma Epidermal Growth Factor Receptor Variant Testing. Ann Lab Med. 2022 Mar 1;42(2):141–9.

28. Wong KHK, Tessier SN, Miyamoto DT, Miller KL, Bookstaver LD, Carey TR, et al. Whole blood stabilization for the microfluidic isolation and molecular characterization of circulating tumor cells. Nat Commun. 2017 Nov 23;8(1):1733.

29. Lippi G, Salvagno GL, Montagnana M, Franchini M, Guidi GC. Venous stasis and routine hematologic testing. Clin Lab Haematol. 2006 Oct;28(5):332–7.

30. Doucet M, Becker KF, Björkman J, Bonnet J, Clément B, Daidone MG, et al. Quality Matters: 2016 Annual Conference of the National Infrastructures for Biobanking. Biopreserv Biobank. 2017 Jun;15(3):270–6.

31. Ntzifa A, Lianidou E. Pre-analytical conditions and implementation of quality control steps in liquid biopsy analysis. Crit Rev Clin Lab Sci. 2023 Dec;60(8):573–94.

